# Mental Health and Service Utilization Among Asian-Indians in the United States: A Scoping Review

**DOI:** 10.1101/2024.12.19.24319344

**Authors:** Rose Mary Xavier, Harjas Kaur, Ram N. Kalidindi

**Affiliations:** School of Nursing, University of North Carolina at Chapel Hill, NC; Gillings School of Public Health, University of North Carolina at Chapel Hill, NC; College of Arts and Sciences, University of North Carolina at Chapel Hill, NC

**Keywords:** Asian Indians, mental health, mental health disorders, mental health service utilization, disparities

## Abstract

**Background:** Asian-Indians living in the US are severely underrepresented in mental health research. Though there are more studies on Asian-Indian mental health now than there was a decade ago, much remains unknown including the experience and prevalence of mental health disorders, service utilization, impact of cultural influences, and barriers and facilitators in seeking mental health support and treatment. The purpose of this scoping review was to examine mental health and mental health service utilization among Asian-Indians in the US.

**Methods:** We conducted a scoping review of published literature. Two reviewers independently screened 2687 articles for potential inclusion—1976 were excluded based on title and abstract review alone and 675 on full-text review. An additional 8 articles identified during full text review resulted in a total of 44 articles included in the scoping review.

**Results:** Of the 44 studies, 33 used a quantitative design, 3 studies used a multiple or mixed methods design and 8 used a qualitative design. Given the broad theme of our research question and since the final list of studies included varied widely in their focus and methods, we present our results using a descriptive, narrative synthesis.

**Conclusions:** Cultural values, familial roles and support, and stigma significantly impact both mental health and help seeking behaviors of Asian Indians. There are major gaps in the literature surrounding child and youth mental health, mental health disorders and pathways to mental health care. These findings are discussed in the context of relevance for practice, policy and the need for high quality research.

## Introduction

Studies have shown that most ethnocultural groups in the United States (US) including Asian Americans, are less likely to utilize mental health resources, even though they may have a greater need for doing so [10]. Asian Americans are the least likely to utilize mental health services and report the lowest prevalence of mental health disorders when compared to other racial and ethnocultural groups in the US [56, 60]. The lower prevalence of mental health disorders is believed to be due to underreporting from cultural stigma related to mental health disorders which subsequently creates barriers to accessing mental health services [56, 60].

Stereotyped as the “model minority” for academic and economic success than other ethnocultural groups, Asian Americans are also attributed with perseverance implying “greater mental and emotional resiliency” leading to perceived reduced need for mental health services [3, 13, 60].

Most mental health research on the Asian Americans consider Asian Americans as a monolith, despite differences in the country of origin and cultural influences which can influence mental health, development of mental health disorders and mental health service utilization [30].

Asian Indians are a rapidly growing population in the US, doubling in numbers in the last two decades with an estimated 4.9 million US residents reporting Asian Indian ancestry of origin [20]. Among the six largest Asian origin groups in the US—Chinese, Indian, Filipino, Vietnamese, Korean and Japanese—Asian Indians are the second largest accounting for 21% of the Asian origin group [29]. A significant proportion (68%) of Asian Indians residing in the US are foreign born [29] immigrating to the US as high-skilled workers, for career and academic opportunities and their family members (e.g., aging parents) joining them later [39]. Asian-Indians as an Asian sub-ethnic group have distinct social and cultural norms such as family-oriented and collectivistic values, emphasis on religion and spirituality and a strongly patriarchal society [57].

Culture can shape expression of mental health symptoms, illness and help seeking behaviors. The distinct cultural values of Asian Indians and the effects of immigration are important factors that can exert unique effects on the mental health of Asian Indians including seeking mental health resources and services [4]. Though there are more studies on Asian-Indian mental health now than there was a decade ago, much remains unknown including prevalence and service utilization, impacts of cultural influences and stigma as well as the barriers and facilitators in seeking mental health support and treatment. In the context of a rapidly growing population and by the virtue of the status of Asian Indians as immigrants, combined with the rising proportions of US born Asian Indians, we focus this scoping review on factors influencing mental health and mental health disorders as well as mental health service utilization among Asian-Indians in the US.

## Methods

We conducted a scoping review of primary source literature on mental health and mental health service utilization among Asian-Indians in the US. With the assistance of a trained health sciences librarian, we identified and searched the databases PubMed and PsycINFO using MeSH terms “Asian-Indians,” “Asian-Indian immigrants,” or “Asian-Indians in the US,” “mental health”, “mental health disorders,” “mental health services,” “United States,” and “India/ethnology” to identify articles published until June 18, 2024. A complete list of search terms is provided in the supplement S1. We did not restrict by publication dates and thesis/dissertations were also included. To meet criteria for inclusion in our review, Asian-Indians had to be the population focus OR be included in the study and disaggregated as a subgroup for analysis. We included only those studies that focused either on mental health or mental health service utilization among Asian Indians living in the US. We excluded studies that did not include Asian Indians, reviews, non-research articles and studies that were conducted on Asian Indians living outside the US. Since the purpose of our study was to examine Asian Indian individuals’ mental health and service utilization in the US context, we also excluded studies that primarily focused on comparisons between Asian Indians living in the US *vs* India and other global regions. 2719 articles were identified through the database search and were imported to a review management tool [7] to assess if they met our study criteria for inclusion. After removing 32 duplicates, 2 reviewers independently screened 2687 articles for potential inclusion. 1976 were excluded based on title and abstract review alone and 675 were excluded based on the full-text review (see Fig. 1). During the full text review stage, we identified an additional 8 articles that met our inclusion criteria resulting in a final list of 44 articles which were included in this scoping review.

**FIGURE 1.**
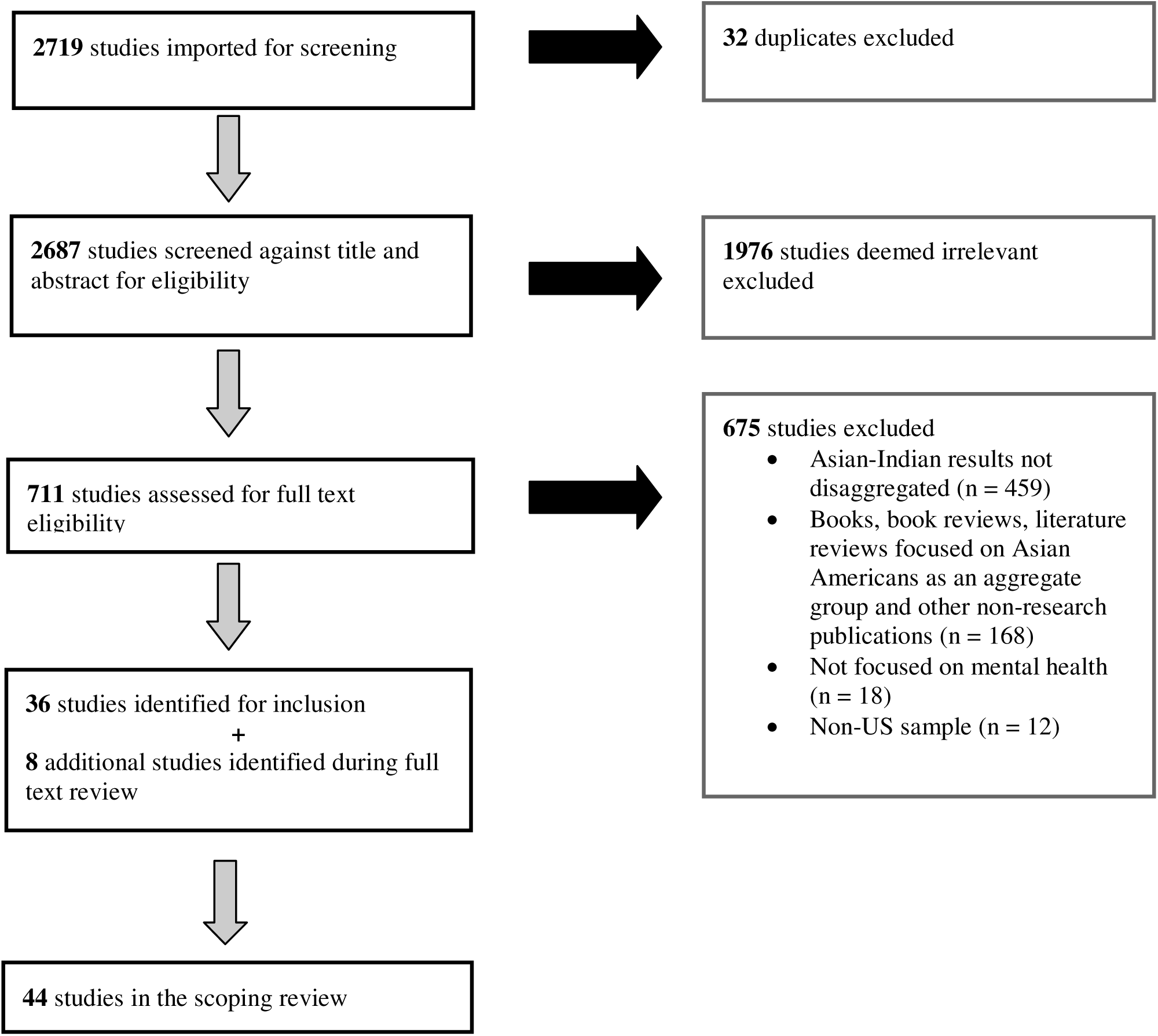
Flowchart outlining article selection process for inclusion in the scoping review.

## Results

Of the 44 studies included in the final analysis, 8 used a qualitative design (Table 1), 33 used a quantitative design (Table 2) and 3 studies used both quantitative and qualitative approaches in a multiple or mixed methods design (Table 3). 19 out of the total 44 studies were dissertations; 15 out of the 44 studies included participants other than Asian Indians. Given the broad theme of our research question and since the final list of studies included varied widely in their focus and design, we present our results using a descriptive, narrative synthesis approach.

**TABLE 1:** Summary of 8 studies using qualitative methods.

| Author(s) | Study design | Aim of study | Study sample description | N (Asian Indian N/Total) | Relevant findings |
| --- | --- | --- | --- | --- | --- |
| **Patel 2003** | Descriptive, narrative discourse | Examine how help seeking attitudes and behaviors impact psychological distress among Asian Indians | Recently immigrated Asian Indian Hindu adults; age 30-50 years; clinical therapy (n=3) and non-clinical (n=3) group | 6/6 | -Relational issues and immigration were major stressors.<br>-Medical referral to therapy was the path to mental health care<br>- Confidentiality a major concern in receiving care |
| **Keeling 2004** | Exploratory descriptive | Explore experiences of a self-identified mental health problem during narrative therapy | Asian Indian women; international students; age 22-30 years | 7/7 | - Self-identified problems were related to cultural adjustment<br>- Narrative therapy techniques were culturally appropriate |
| **Argo 2010** | Exploratory descriptive | Explore the effects of cultural and religious/spiritual variables on experiences of psychotherapy | 1<sup>st</sup> generation Asian Indian Hindu adults—born in India and migrated to the US in youth; in psychotherapy; age 28-60 years | 13/13 | - Favorable view of psychotherapy.<br>- Stigma and privacy concerns<br>- Religiousness and spirituality did not affect views on psychotherapy<br>- Higher acculturation associated with higher number of sessions and a preference for non-Indian therapist. |
| Lee et al., 2011 | Multiple focus groups | Collect data on health needs | Sample from 13 Asian Americans communities; age >18 years | 6/174 | - Mental health among the top three concerns for Asian Indians and other Asian Americans<br>- Need for awareness education on mental health |
| **Jain 2015** | Ethnography and grounded theoretical approach | Explore marital experiences of Asian Indian couples and the implications for culturally competent therapy | Immigrant heterosexual couples; age 22-50 years | 12/12 | - 100% agreed that they would be comfortable accessing mental health services for their spouse and/or children but only 33% would do it for themselves |
| **Sandhu 2016** | Interpretive phenomenology | Explore sociocultural factors, preexisting attitudes and barriers relating to mental health services | Asian Indian Punjabis; 7 community members and 3 mental health professionals; age 29-43 years | 10/10 | - Conflicting traditional and western values<br>- First line of support is family and friends<br>- Stigma, denial, and lack of awareness are barriers<br>- Increasing awareness, resources, anonymous support, group support and physician referrals are needed for service utilization |
| Sharma et al., 2022 | Multiple case study approach | Understand the experiences of Asian Indian Americans when seeking mental health services | 1<sup>st</sup> and 2<sup>nd</sup> generation Asian Indian adults; 20-39 years | 7/7 | - Stigma and stereotypical views of mental health, limited awareness and unfavorable experiences in therapy were major barriers |
| **Matam 2022** | Phenomenology | Examine mental health help seeking following intimate partner violence | Immigrant Asian Indian women; age 25-35 | 10/10 | - Stigma and lack of cultural competence among clinicians were barriers |

*Note:* Bolded author names are dissertations/thesis.

**TABLE 2.** Summaries of 29 studies using quantitative methods.

| **Author(s)** | **Study design** | **Aim of study** | **Study sample description** | **N (Asian Indian /Total)** | **Relevant findings** |
| --- | --- | --- | --- | --- | --- |
| Sodowsky 1991 | Randomized 2-group design | Explore counselor credibility perceptions among Asian and White American students. | University students; age not provided; Included White American, Asian Indian and South Korean groups | 48/129 | -Asian Indians exposed to culturally consistent counseling perceived significantly greater trustworthiness in counseling than Asian Indians exposed to culturally discrepant counselling |
| **Derry 1996** | Cross-sectional survey | Explore the degree and direction of length of US residence and cultural assimilation (retention or rejection of traditional marriage customs) on psychological help seeking behaviors | Indian American adult participants; mean age 42.3 (SD = 14.9); 12/52 participants were Asian Indian psychotherapists | 52/52 | - Cultural assimilation and being a woman but not length of US residence or age associated with amenability to psychotherapy<br>- Despite higher levels of amenability to psychotherapy, 30% report that they did not believe in the efficacy of psychotherapy |
| Roberts et al., 1997 | Cross-sectional survey |  | School based survey of 6-8 grade students in Houston; age 10-17 years old | 188/4186 | - Compared to White Americans, Indian Americans had the lowest lifetime rates for suicidal ideation, thoughts, plan and attempts for past 2 weeks as well as across the lifetime |
| Han & Liu, 2005 | Secondary analysis of Medical Expenditure Panel Survey 1996-2000 | Examine disparities in psychiatric prescription drug use between White, Black, Hispanic and Asian Americans | US adults 18-64 years with one or more diagnoses of mental illness; mean age 41 years | Not provided /4338 | - 19% of sample represent the 3 minority groups<br>- Asian Indians diagnosed were the least likely to use prescription drugs compared to all other racial groups. |
| Goyal et al., 2005 | Quantitative, descriptive | Determine incidence of post-partum depressive symptoms in immigrant Asian Indian women | Asian Indian women between 2weeks to 12 months postpartum and birthed a live, healthy infant; mean age 29 years(SD=3.43) | 58/58 | - 24% screened positive for major post-partum depressive symptoms and 28% for minor post-partum depressive symptoms |
| **Chuang 2005** | Secondary analysis of Mental Health and Mental Retardation Authority of Harris County archival mental health services dataset from 1993-1999 | Compare Asian Indian, Vietnamese and Chinese Americans with regards to utilization rates, diagnoses, and resource use | 655 Asian Americans; age > 18 years | 136/655 | - Depression most prevalent diagnosis across all groups; schizophrenia prevalence was the same as depression among Asian Indians<br>- Compared to other groups, Asian Indians and Chinese Americans used services at the same rate, had greater outpatient services than inpatient services |
| **Nagra 2005** | Cross-sectional; questionnaires were self-administered (sent via mail) | Examine associations of psychological distress and mental health service | 1<sup>st</sup> Asian Indian students; age 18-35 years | 253/253 | - Perceived social support negatively correlated with distress whereas acculturation, ethnic identity, academic distress, and SES were not associated with |
|  | or by an assessor | utilization in Asian Indians |  |  | psychological distress<br>- Acculturation levels, ethnic identity, SES, time spent in the US were associated with willingness to seek mental health service |
| Huang et al., 2007 | Secondary analysis of Early Childhood Longitudinal Survey-Birth Cohort | Examine prevalence of maternal depression and help seeking patterns | Mothers, age >15 years, with children born in the US in 2001 | 205/8462 | - Foreign born Asian mothers including Asian Indians have a higher prevalence of depressive symptoms than US-born Asian mothers<br>- Small sample so ethnicity related help seeking patterns not examined |
| **Sandil 2009** | Cross-sectional survey | Examine Asian Indian women's counseling and counselor expectations | Asian Indian women; age not provided | 149/149 | - Level of enculturation was associated with counselor preferences. |
| **Mohan 2011** | Cross-sectional survey | Examine acculturation effects on attitudes towards help-seeking for children among Asian Indian parents or caregivers | Asian Indian parents or caregivers (for at least 6 months) of children 7-17 years, resided in the US for a minimum of 1 year; from 3 states in the US; age 31-60 | 73/73 | - Acculturation was not associated with openness, stigma, or intention to seek mental health services for children |
| Wong et al., 2014 | Secondary analysis of National Epidemiologic Survey of Alcohol and Related Conditions | Examine association of proportion of life in the US and lifetime suicide ideation | US Asian Americans; mean age 41.2 (range 18-94) | 187/1332 | - Indian Americans report the lowest rates for suicidal ideation compared to all other Asian Americans<br>- Association of the proportion of life lived in the US with suicidal ideation was the strongest for Indian Americans |
| Aratani & Liu, 2015 | Secondary analysis of administrative data from 2001-2006 | Examine association of English proficiency and ethnicity on mental health services discontinuation | Children <18 years utilizing county mental health services from 11 California counties | 3%/59218 | - English and non-English speaking Asian Indians were no more likely to drop out of mental health services compared to White Americans; comparisons against other Asian sub-ethnicities not reported |
| Lane et al., 2016 | Cross-sectional survey | Examine suicidal ideation and ethnic predictors among South Asian American groups | Undergraduate students, mean age 18.5 years, SD=0.93 | 67/204 | - Asian Indian young adults have higher levels of suicidal ideation compared to Bangladeshi and Pakistani groups<br>- Hopelessness moderated the association between suicidal ideation and South Asian ethnic group. |
| Nadimpalli et al., 2016 | Secondary analysis of the Mediators of Atherosclerosis in South Asians Living in America study | Examine relationship between discrimination and mental health (depressive symptoms, anger and anxiety) | South Asians in the US; age 40-83 years | 733/906 | - Higher discrimination levels significantly associated with higher depression, anger and anxiety<br>- Stronger cultural beliefs moderate the association between discrimination and depression<br>- Active coping style moderates the relationship between discrimination and anger |
| Turner & Mohan, 2016 | Cross-sectional survey | Explore parents past use of services, stigma and attitudes influence intentions to use child mental health services | Asian Indian parents of children ages 6-17 years from 5 states; Mean age 42.33 (SD=6.19) | 89/89 | - Parents past use of mental health services correlated with more positive help seeking attitudes but not help seeking intentions<br>- Parental attitudes and not stigma associated with a greater likelihood of seeking child mental health services |
| Roberts et al., 2016 | Cross-sectional survey | Explore sociocultural factors related to depression in Sikh immigrants | Asian Indians; mean age 41.9(SD=15.4); 13% US born | 350/350 | - Negative religious coping and anxiety associated with depressive symptoms |
| **Narikkattu 2018** | Cross-sectional survey | Examine association of religiosity and attitudes towards help seeking behavior in Christian Indian Americans | Christian Indian Americans recruited from a University Asian American cultural center; age 18-63 years; 78% of participants US born | 93/93 | - Religiosity negatively correlated and acculturation positively correlated with attitudes towards help-seeking |
| Jang et al., 2019 | Secondary analysis of Asian American Quality of Life survey | Explore the unmet need for mental health care | Asian Americans; >18 years old | 574/2609 | - Asian Indians had the second highest odds of using mental health services.<br>- Examined service use in severe mental illness, but sample sizes are very small for meaningful interpretation |
| **Panjwani 2020** | Cross-sectional survey | Examine associations of immigration status and US length of stay on quality of life and depression among Asian Indians | Asian Indians, mean age 37.6 years (SD=8.4 years) | 73/73 | - Immigration status is associated with quality of life but not depressive or anxiety symptoms with Asian Indians with non-immigrant status reporting the lowest quality of life. |
| **Thomas 2020** | Cross-sectional survey | Understand help-seeking behaviors and their relationship to psychological resilience in Asian Americans | 1<sup>st</sup> or 2<sup>nd</sup> generation immigrant Asian-Indian; >18 years old | 106/106 | - ~ 20% had used mental health services specifically 'personal' and marriage counseling<br>- Resilience was not associated with mental health help-seeking behavior |
| DeVitre & Pan, 2020 | Cross-sectional survey | Explore the relationship between internalized values and attitudes towards mental health services | 1<sup>st</sup>, 2<sup>nd</sup> or 3<sup>rd</sup> Asian Americans; included Chinese Americans and Indian Americans | 49/75 | - Enculturation (adherence to heritage cultural values) had a negative mediating effect and acculturation (adherence to European American values) had a positive mediating effect on the relationship between attitudes towards mental health and help seeking |
| **Mistry 2021** | Cross-sectional, survey | Examine mental health help-seeking behaviors in Indian Americans | Asian Indians; mean age 42.7 (SD=22.8) | 95/95 | - Higher acculturation positively associated with help seeking behaviors<br>- Age was negatively correlated with help seeking |
| Jin et al., 2022 | Cross-sectional survey | Examine ethnic cultural values and mental health among Indians | Asian Indians >18 years old; mean age 34.7 | 231/231 | - Three distinct subgroups each with variations in symptom distress, barriers to accessing care and treatment preferences were identified on ethnic cultural values |
| Daulat & Wadhwa, 2022 | Cross-sectional survey | Examine Indian American women's dual cultural identity and its impact on identity-self acceptance, perceived workplace diversity climate and work engagement | Indian American women >18 years of age, who have lived in the US since age 8 or before | 99/99 | - Mental health mediates the relationship between identity-self acceptance and work engagement. |
| DeVitre et al., 2022 | Cross-sectional survey | Explore Indian American undergraduates' psychological wellbeing using a psychosociocultural perspective | 1<sup>st</sup> generation Indian American university undergraduates; mean age 19.9 (SD=1.58) | 122/122 | - Seeking professional care was the least used coping approach; planned action by themselves followed by "talking to others" was most frequently used. |
| Balaraman et al., 2022 | Secondary analysis of National Health Interview Survey 2006-2018 | Examine prevalence of psychological distress and mental health service utilization in Asian American adults compared to their Non-Hispanic White (NHW) counterparts matched to age, sex and socioeconomic status | Nationally representative US sample; Asian Indian, Chinese, Filipino and NHWs included in the analysis; age >18 years | 4242/282,382 | - Mental health service utilization is lowest for Asian Indians (2.8%); No increase in service utilization despite increase in psychological distress from 2006 to 2018.<br>- Lowest odds of service utilization (adjusted odds ratio 0.51) compared to other groups after adjusting for age, sex, race, education, income, marital and nativity status |
| **Patel 2022** | Observational | Explore perceptions of mental health and help-seeking attitudes among Asian Indians Americans | Asian Indian Americans; mean age 41 (range 18-89); women (n=75); first-generation (n=67) | 120/120 | - No significant differences between first-generation and second-generation Asian Indian American's perceptions on mental health and help-seeking behavior.<br>- Younger participants had more positive mental health views.<br>- Women had more positive perceptions toward mental health services and are more likely to seek help.<br>- Acculturation not a significant predictor for attitudes related to mental health. |
| Siddiqui & Sambamoorthi, 2022 | Secondary analysis of the National Health Interview Survey 2012-2017 | Examine psychological distress among Asian Indians and report comparisons against Non-Hispanic White sample | Nationally representative US sample; age > 18 years | 2218/124,617 | - Asian Indians had a lower prevalence of psychological distress compared to Non-Hispanic Whites |
| **Sundaram 2022** | Randomized control trial | Assess the perceived utility and satisfaction of different versions (genetic, culturally adapted, and customized) of | Asian Indians living in the US and above the age of 18 (N=105). Average age of 31.84 years (range 18-63); 68% | 105/105 | - Higher degree of perceived utility and satisfaction for the culturally adapted and customized versions of the MITRE.<br>- Higher mood ratings associated with greater perceived utility for MITRE. |
|  |  | MITRE (Micro Interventions and Tools for Reducing Emotional distress) among Indians living in the US. | females; 60% single; 57% had a bachelor's degree or above |  |  |
| Ikram et al., 2023 | Cross-sectional online survey | Determine predictors of COVID-19 related mental health among Asian Indians | Asian Indian adults; age 18-96 | 289/289 | - 46% of the study sample reported poor mental health; 20% reported sleep disturbance<br>- Sleep disturbance and age were the strongest predictors of poor mental health<br>- Higher income levels associated with poor mental health |
| **Savani 2023** | Cross-sectional survey | Examine relationship between acculturation and psychological help seeking among South Asian women | 2<sup>nd</sup> generation Asian Indian women living in the United States; mean age =31 (range 18-60) | 89/89 | - Acculturation levels not associated with help-seeking attitudes. |
| Compton 2023 | Cross-sectional | Examine the associations between PTSD symptom severity and institutional/ internal help-seeking barriers for mental health. | Trauma-exposed Indians living in the southwest of the United States; mean age of 31.61 years; 71.4% women (n=55); First generation (n=50); Second generation (n=26). | 77/77 | - Marginal association between PTSD symptom severity and institutional barriers.<br>- Ageist attitudes correlated with lower PTSD symptom severity.<br>- Difficulties in transportation associated with higher PTSD symptom severity |
| DeVitre 2024 | Cross-sectional | Explores how the internalization of the 'Model Minority Myth' in a university environment affects Indian Americans Students' perception of mental health services. | Indian American undergraduates; ~50% women; mean age of 21 years | 205/205 | - University setting impacted the internalization of the Model Minority Myth and led to increased pressure and expectation for Indian American students to succeed without the need for mental health services<br>- Decreased psychological well-being and less desirable attitudes toward seeking mental health services. |

*Note:* Bolded author names are dissertations/thesis.

**TABLE 3.** Summaries of 3 studies using multiple or mixed methods.

| **Author(s)** | **Study design** | **Aim of study** | **Study sample description** | **Asian Indian N/Total** | **Relevant findings** |
| --- | --- | --- | --- | --- | --- |
| **Joseph 1994** | 2-group design; questionnaires and participant interviews | Examine ethnic identification to understand psychotherapy evaluation in African vs Asian Americans | African and Asian Indian college students; mean age 20.7 years | 27/92 | - Ethnic identification stages impact levels of personal and collective self-esteem for ethnic minority individuals as well as evaluation of psychotherapy treatment approaches but these do not differ by ethnicity |
| De Gagne et al., 2015 | Concurrent triangulation mixed methods- survey and focus groups | Evaluate the health care experience of Asian Indians | Asian Indian immigrants; 40-64 years; | Survey N 125/125. Focus group N = 10 | - 94.4% rated mental health good, 5.6% rated it as neutral; None rated mental health as poor in the survey despite focus group participants discussing psychosocial stress |
| Joseph et al., 2020 | Survey which included open-ended questions | Examine acculturation and mental health among Asian Indian women | 1<sup>st</sup> (N=42) or 2<sup>nd</sup> (N=31) generation immigrant Asian Indian Malayalee (from the Indian state of Kerala) women; mean age 28 (range = 16-83) | 73/73 | - Attitudinal distancing from heritage culture associated with greater depression symptoms |

*Note:* Bolded author names are dissertations/thesis.

### Recurring themes in qualitative studies

Eight qualitative studies were meta-synthesized, and results are reported below.

#### Help seeking behaviors

Six of the 8 qualitative studies discussed help seeking behaviors among Asian-Indians, including coping mechanisms, methods of managing mental health, and how often and what type of mental health care they seek [2, 22, 33, 42, 45, 50]. Two studies reported that participants were more likely to obtain help for their loved one than get help for themselves for mental health issues [22, 45]. Sandhu (2016) reported that none of the study participants mentioned seeking professional mental health care as a personal option for help seeking; most participants reported speaking with family and friends as a form of help seeking and that they would only consider seeking professional mental health care if other options failed [45]. Other studies reported similar findings with families providing direct or indirect mental health support and participants stating they would seek professional help only if their problems couldn’t be resolved informally on their own or by family support [42]. Participants also reported a mistrust of professional mental health care, as well as not knowing enough about seeking mental health care to do so [50]. In addition, participants often avoided help-seeking from professionals because it would be more socially acceptable to seek help from family and friends [42]. Another common reason for not seeking help was stigma and mental health issues being taboo within the community [2, 33, 45, 50]. Job status also influenced help seeking behavior among Asian-Indians by providing adequate benefits and more financial resources to access professional care [2]. Participants also reported that they would only want to seek mental health care if they were able to see a therapist who is either Asian-Indian or familiar with Asian-Indian culture and reported struggles in finding a therapist meeting these criteria [33].

#### Stigma

Stigma surrounding mental health and professional mental health care was another main theme in many studies [2, 33, 45, 50]. Cultural stigma around seeking psychological help for mental disorders was strong amongst participants and although many felt they would be comfortable receiving therapy, they reported not feel comfortable letting their family members know [2]. Strong negative attitudes towards psychotherapy within their families, and an upholding of traditional Indian values of keeping matters private and hidden were reported by participants [2]. In addition, one study reported that participants felt guilty for speaking about their mental health struggles out of fear of negatively portraying their family [50]. Stigma also influenced recruitment to studies where many people who inquired about the study were not inclined to participate due to stigma around mental illness and psychotherapy and the idea that mental illness does not really “exist in Indian culture” [2, 50]. Stigma significantly hindered help-seeking attitudes, a strong theme in qualitative studies included in our synthesis.

#### Role of the family

Family is often the main source of mental health support for Asian-Indians. Of the 8 qualitative studies, 3 studies identified the role families played in providing support [33, 42, 45]. In one study for example, all participants stated that their families played an important role in support for their mental health (Patel, 2003). Five out of the 7 participants in another study stated that they would seek help from their families for their mental health needs (Sandhu, 2016). Some participants reported that they couldn’t find the support they needed for mental health issues from their families because mental health was never discussed in their households (Matam, 2022). For others, relying on their family for coping is not an option because struggling with mental health is seen as a sign of weakness by their families [50].

#### Cultural factors

Cultural factors that prevent Asian Indians from seeking out mental health services was another major theme amongst qualitative studies. In one study, participants reported experiencing conflicting cultural values, preexisting thoughts and attitudes about mental health problems, and resistance to seeking help regarding mental health problems (Sandhu, 2016). The traditional value of keeping matters private influenced negative perceptions regarding psychotherapy, which contributed to stigma and an overall resistance and lack of help-seeking attitudes [2, 45]. Judgment from the Indian community if one openly received psychological help, was another major inhibiting factor [45]. In another study, participants described feeling held back from seeking mental health care because certain cultural values made them feel like speaking out about their mental health would cause people to view them as “crazy” [50].

Participants in another study reported not feeling supported by their cultural community as seeking mental health treatment was considered a taboo [33].

#### Need for mental health awareness

Mental health awareness was another major theme across qualitative studies, with 5 studies reporting that an increased awareness regarding mental health services and mental illness was needed and would increase utilization and that a lack of awareness inhibited service utilization [2, 22, 32, 45, 50]. There appeared to be a consensus among these studies that lack of awareness was a big factor inhibiting the Asian Indian community from seeking out mental health services. These studies also generally identified a need for increased availability of mental health resources, the need for increased awareness of existing resources as well as resources on how to access them. Studies reported that mental health awareness could be improved through educational [32], informational events that could be delivered through religious, community organizations [45].

### Central themes in quantitative studies

Of the 36 studies (33 quantitative and 3 mixed/multiple methods) that employed a quantitative design, we identified the following overarching themes.

### Mental health and help seeking

#### Acculturation and cultural factors

Acculturation and the impact of cultural factors on mental health and help seeking was the most studied concept with 14 out of the 36 quantitative studies examining the influences of acculturation either on mental health and/or help seeking behaviors among their participants [8, 11, 12, 14, 24, 26, 34, 35, 38, 40, 41, 46, 47, 53]. Three of these studies were focused on Asian Indian women [26, 46, 47]. Cultural factors and acculturation play a significant role in mental health and help seeking among Asian Indians.

Pressure to adhere to cultural values was associated with higher levels of psychological distress [24]. Asian Indian university students were reported to have a greater trust in culturally consistent counseling [52]. Culturally adapted interventions also result in higher perceived utility and satisfaction [53].

Studies generally defined acculturation as the ability to successfully navigate two different cultures and these studies reported equivocal findings with some reporting no association [47] whereas others reporting a positive association [40]. Please see Table 4 for the various scales used to measure acculturation and help seeking across quantitative studies.

**TABLE 4.**
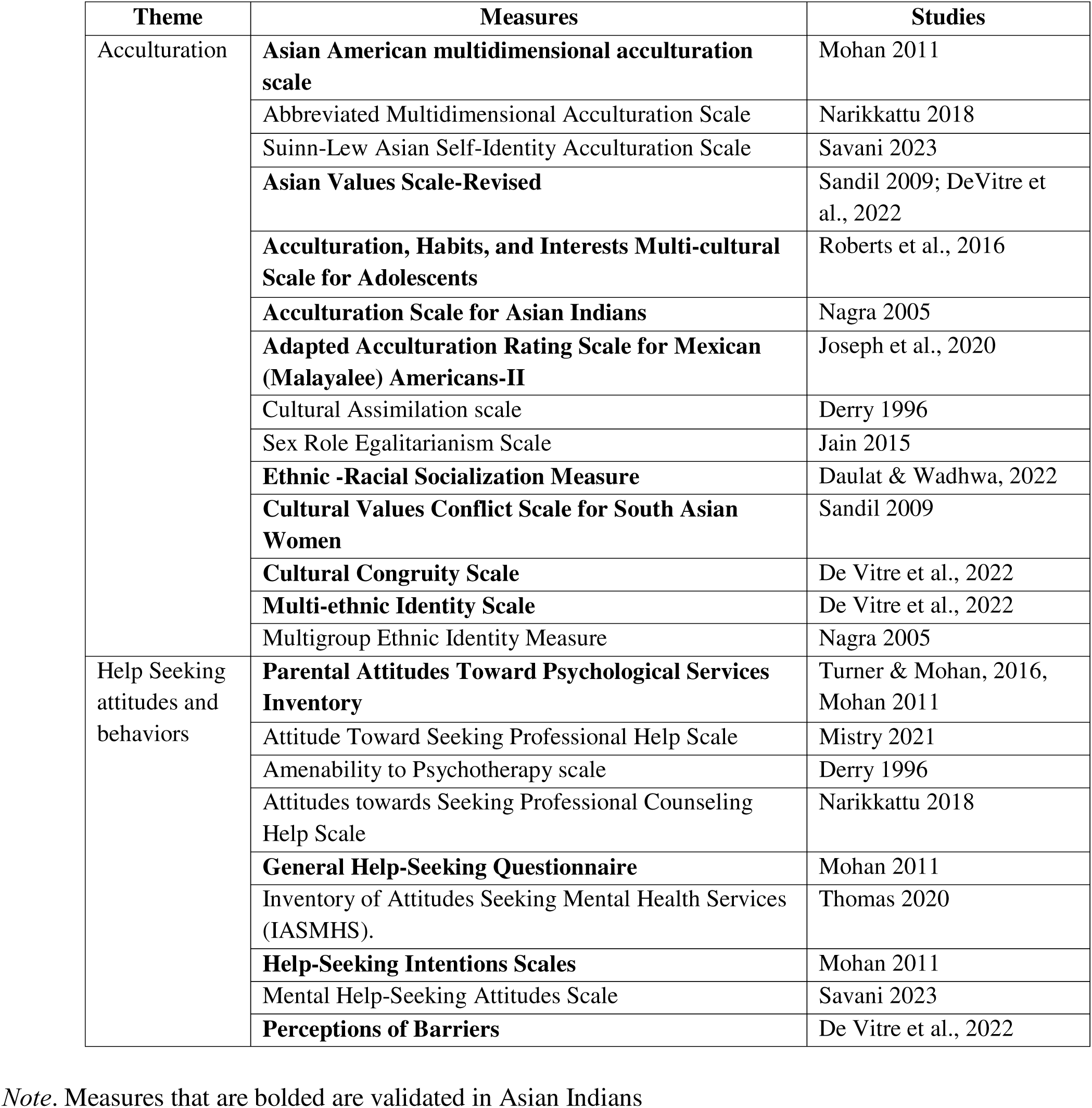
Measures used across studies to assess most frequently studied concepts.

Acculturation, defined as adherence to European American values, was reported to have a positive mediating effect on relationship between attitudes towards mental health and help seeking [14]. Though acculturation was not associated with intent to seek mental health services [35], it was generally positively associated with willingness to obtain help [38], help seeking behaviors [34] amenability to psychotherapy [11] and counseling style and counselor preferences [46]. Asian Indians who are less acculturated also face more barriers, both internal and external, towards mental health help seeking [24].

Ethnic identity, the extent to which an Asian Indian person self-identifies to be part of the Asian Indian ethnic group, also influences mental health and help seeking. For example the more Asian Indian women attitudinally distanced themselves from their heritage culture, the more depressive symptoms they experience [26]. Jin et al (2022) reported on three distinct subgroups each with variations in symptom distress, barriers to accessing care and treatment preferences identified on ethnic cultural values. Savani (2023) focused on the relationship between cultural identity self-acceptance and work engagement, reporting a significant mediating effect of mental health between the two. Ethnic identity though not associated with psychological distress was reported to be associated with willingness to seek mental health service [38].

#### Socio-demographic factors

Nine out of the 36 quantitative studies reported on the influences of socio-demographic factors such as age, gender and socioeconomic status (SES) on mental health and help seeking [3, 11, 22, 23, 26, 34, 38, 40, 55]. Three studies reported statistically significant differences between older and younger individuals with regards to both mental health and help seeking attitudes with younger individuals more willing to seek mental health care [3, 26, 34]. Two studies reported that age related differences were not statistically significant in their sample [40, 55] and one study reported a negative correlation between age and amenability to psychotherapy [11]. Help seeking among Asian-Indians over the age of 50 was reported to be influenced by factors such as financial stability, cultural barriers, English language proficiency and not prioritizing mental health concerns [34]. Women were more open to seeking mental health services and more amenable to psychotherapy [11]. One study reported that married and older women had better self-esteem and psychological outcomes than younger women [22].

Other sociodemographic factors that reported a positive association with willingness to seek mental health service were high SES [22, 38] and longer length of stay in the US [38].

Paradoxically, during the COVID-19 pandemic higher income was associated with poor mental health [19]. Religiosity negatively correlated with attitudes towards help seeking [40] and unsurprisingly increased levels of social support from friends, relatives, and other people were associated with lower levels of psychological distress and a lower likelihood of seeking out mental health services [22].

### Mental health symptoms, prevalence, and service utilization

Thirteen out of the 36 quantitative studies examined clinically important parameters such as prevalence of mental health symptoms, psychological distress, mental health disorders and mental health service utilization [1, 3, 5, 6, 16–19, 23, 31, 37, 44, 59]. Of these, four examined mental health service utilization [1, 3, 17, 23], 2 studies examined maternal depressive symptoms [16, 18], 3 studies examined suicidality [31, 44, 59] and one study examined help-seeking barriers and PTSD symptom severity [6].

Using county mental health administrative records, Chuang (2005) reported that depression was the most prevalent mental health diagnosis among all Asian subethnic groups, but Asian Indians had an equal prevalence of depression and schizophrenia [5]. A study examining the prevalence, maternal depression used the Early Childhood Longitudinal Survey-Birth Cohort reported that foreign born Asian mothers including Asian Indian mothers had a higher prevalence of depressive symptoms than their US born counterparts [18]. Goyal and colleagues [16] in a convenience sample of 58 women, reported that 24% of the sample screened positive for major post-partum depressive symptoms and 28% for minor post-partum depressive symptoms.

Data on suicidality was inconsistent. In regional samples, Asian Indian middle school children were reported to have the lowest lifetime rates as well as the immediate past 2 weeks for suicidal ideations, thoughts, plans and attempts [44]. In contrast, Asian Indian young adults (mean age 18.4 years) have higher levels of suicidal ideation when compared to other South Asian groups with hopelessness significantly increasing the risk [31]. In a nationally representative sample, Indian Americans were reported to have the lowest odds of suicidal ideation, but the strongest odds when accounting for the moderating effects of proportion of years lived in the US [59].

Reports on mental health service utilization was inconsistent. In a nationally representative US sample extracted from the National Health Interview Survey, Balaraman et al. (2022) reported that service utilization was lowest for Asian Indians (2.8%) despite significant increases in psychological distress between 2006 to 2018; Asian Indians had the lowest odds of service utilization (adjusted odds ratio 0.51) compared to other Asian groups and other racial groups after adjusting for age, sex, race, education, income, marital and nativity status [3]. These findings contrasted an earlier report [23] based on regional data that Asian Indians had the second highest odds of using mental health services [23]. Aratani & Liu (2015) reported that both English and non-English speaking Asian Indians were no more likely to drop out of mental health services for children when compared to White Americans, though they did not report comparisons with other Asian subethnicities. Asian Indians were also reported to be the least likely to use psychiatric prescription drugs compared to White, Black, Hispanic, and other Asian subethnic groups [17].

## Discussion

Our scoping review identified several themes related to the mental health experiences of Asian Indians, including mental health help-seeking behaviors, stigma, family involvement, cultural influences as well as the pressing need for increased mental health awareness and culturally sensitive pathways to access mental healthcare resources.

Asian Indians are a rapidly growing community and the second largest immigrant group in the US. The Asian Indian community is a mosaic of recent immigrants and long-term residents including a growing proportion of US born individuals. The Asian Indian demographic is far from a monolith, yet recent surveys show certain trends at the national level. For instance, roughly 75% of Asian Indians reportedly hold a bachelor’s degree, presenting the highest median annual household income and lowest poverty rates in the nation [21]. However, this overarching view masks significant intra-group disparities and display variations in aspects such as socioeconomic status [15, 28, 54] and communication abilities [49], which significantly impact mental health, mental health care resource availability and accessibility [3, 19, 38].

The Asian Indian community is characterized by distinctive cultural and sub-cultural values that set them apart. One key attribute is the prevalence of the collectivistic structure exemplified in the traditional joint family structure. This structure where multiple generations cohabit and contribute to a tight-knit familial network, fosters deep ties of support, cooperation, and interdependence among family members [48]. Though Asian Indians in the US are unlikely to have a joint family structure, the collectivistic structure of self-identity that contrasts with the individualistic Western culture has implications for mental health [25, 45] as well as where, how and for who care is accessed [22, 58].

Our review sheds light on the preference of Asian Indians to lean on family support rather than professional mental health services [12, 45], primarily from stigma attached to mental health issues [2, 33, 35, 45, 50, 58] and a lack of knowledge about the available resources to access mental health care [32, 45, 50]. Stigma can impact discrepant or underreporting of mental health symptoms and wellbeing [9]. Mental health discussions are still considered taboo in many spaces, underscoring the influence of cultural norms on mental health experiences.

Insights from the 2020 Indian American Attitudes Survey, which delved into the social experiences of approximately 1200 Asian Indians, highlighted several challenges the community faces with matters of social and ethnic identity, as well as the significant discrimination they often encounter socially, particularly those relating to skin color [51] . It also underscored the apparent homogeneity within social networks and the central role that religion plays within this community pointing to pertinent cultural complexities [51]. Religious beliefs form an integral part of the cultural identity of Asian Indians, influencing lifestyle choices, rituals, and worldview. These cultural norms and values not only shape the day-to-day lives but also significantly impact mental health needs, symptoms, perceptions, help-seeking behaviors and response to interventions [2, 14, 24–27, 35, 37, 38, 45, 52, 53].

Various factors including acculturation and a strong ethnic identity, intertwine intricately to shape the mental health landscape among Asian Indians in the US. The ability to navigate between heritage culture and American culture [14, 45] rather than acculturation [2, 35, 38, 47] positively influences help-seeking attitudes among Asian Indians. Stricter adherence to heritage cultural values is reported to have a negative impact on help-seeking, but transitioning away from heritage culture often parallels an increase in mental health symptoms [26, 37]. Similarly, religiosity is observed to negatively correlate with attitudes towards help-seeking [40], but it is also protective for mental health [43]. Further complexity is introduced by socio-demographic characteristics which influence help-seeking attitudes [3, 11, 19, 34]. These complexities call for tailored interventions that consider the nuances of factors like enculturation, acculturation, and the role of religious and socio-cultural factors in shaping mental health perceptions and help-seeking behaviors among Asian Indians.

Understanding pathways to receiving care is a crucial component in addressing the complexities of mental healthcare utilization within the Asian Indian population. There is a paucity of studies that have examined this and the few studies that addressed this identify that professional care is the least preferred method for addressing mental health challenges [12, 45]. Physician referrals are often needed for mental healthcare utilization [45] and medical referral is often the pathway for accessing counseling and therapy [42] for many Asian Indians. It is noteworthy that despite the increased psychological distress reported by Asian Indians, they are the least likely to utilize mental health services among ethnic groups [3]. This may stem from cultural stereotypes regarding mental health, lack of awareness about available services and/or potential stigma tied to seeking professional help and underscores the urgent need for culturally appropriate services and increased mental health awareness.

### Major gaps and implications

Our review identified major gaps in our understanding of the mental health landscape of Asian Indians in the US. The following are the key areas where more research is urgently needed. *First,* the dearth of epidemiological data pertaining to the prevalence of mental health disorders among the Asian Indian population in the United States is a significant concern. Asian Indians are often lumped into the broader category of Asian Americans and as such existing data does not provide a comprehensive understanding of the full spectrum and complexity of mental health issues faced by this population. An incomplete picture of the mental health landscape can pose challenges to develop targeted and culturally sensitive strategies and there is a compelling necessity for a substantial increase in high-quality, methodologically rigorous empirical research in this population. *Second,* there is a lack of understanding of the impact of the immigration process on mental health. Immigrants face unique stresses and potential traumas that can lead to or worsen mental health issues, but the specific experiences of Asian Indians in the US are underexplored. *Third,* the role of discrimination in mental health outcomes is another area where more knowledge is needed. A recent national survey reported that one in two Asian Indians experience discrimination with ∼30% of the survey respondents reporting discrimination based on skin color [51]. It is known that discrimination can significantly impact mental health, but the experiences of discrimination faced by Asian Indians and the subsequent mental health impact largely remains unknown. *Fourth*, child and youth mental health constitutes another severely understudied aspect. There is a vital need to invest more in research that advances the understanding of child and youth mental health within the Asian Indian population. Given the distinctive socio-cultural factors and concerns such as school issues, peer pressure, and identity formation affecting this group’s mental health, dedicated research is urgently required. *Fifth*, there are few to no studies on many mental health disorders including severe mental illnesses in the Asian Indian population. Research prioritizing psychiatric and mental health disorders are critically needed as these disorders significantly impact individuals’ quality of life and socio-economic potential, making it crucial to better understand their trends, root causes, and effective treatment strategies within the context of cultural factors. *Finally*, pathways to receiving mental healthcare including accessibility and affordability remain critically under-researched. Further study is required to ascertain how Asian Indians navigate mental healthcare systems and the barriers they currently face in accessing mental health care. Design of mental healthcare pathways must be premised on the unique cultural strengths of the Asian Indian population. Existing care models do not sufficiently factor in the unique cultural nuances, strengths, and challenges of this group, necessitating the creation of personalized and culturally sensitive pathways.

Our review has limitations. We focused on mental health and service utilization and though we applied mental health in a broader context, areas that impact mental health such as intimate partner violence and substance use/dependence were not evaluated in our review. Our scoping review aimed for recognizing broad themes or factors informing mental health and service utilization and to identify current knowledge deficiencies for directing specific areas of future research. As such we do not offer a meticulous appraisal of the evidence available [36].

To conclude, our review identified the glaring gaps in our understanding of the mental health landscape of Asian Indians in the US. There is a need for culturally appropriate mental health support systems, resources, and interventions. Building a mental health support infrastructure that resonates with the cultural ethos of the growing Asian Indian population can enhance access to care, early identification of clinical conditions, and adherence to treatment plans, thereby improving overall mental health outcomes.

## Data Availability

This is a scoping review and only published studies were included in the review

## Acknowledgements

We extend our gratitude to Ms. Aditi Nambiar and Ms. Deepa Ramesh for their contributions to article screening and data extraction in the earlier phases of the project.

## Author Contributions

RMX: Conceptualization, Methodology, Investigation, Supervision, Project Administration, Resources, Writing – Original draft, Review & Editing

HK: Data Curation, Investigation, Writing – Original Draft, Writing – Review & Editing RK: Data Curation, Investigation

