## Supplement for "Mental Health and Service Utilization Among Asian-Indians in the United States: A Scoping Review"

**Supplementary material S1. Keywords used for database search**

**PUBMED**

("Mental Health Services"[Mesh] OR "Mental Health Services"[TIAB] or "Mental Health Service"[TIAB]) AND ("Asian Americans"[Mesh] OR "Asian-American"[TIAB] OR "Asian Americans" [TIAB] OR "Asian Indian American"[TIAB] OR "Asian Indian Americans"[TIAB] OR ((Asians [TIAB] or "Asian Indians" [TIAB] OR Asian [TIAB] or "Asian Indian" [TIAB] OR "India/ethnology"[Mesh]) AND ("United States" [TIAB] OR "United States"[Mesh])))

**PsycINFO**

(DE "Mental Health Services" OR DE "Community Mental Health Services" OR DE "Psychological First Aid" OR "Mental Health Services" or "Mental Health Service") AND ("Asian-American" OR "Asian Americans" OR "Asian Indian American" OR "Asian Indian Americans" OR ((DE "Asians" OR Asians or "Asian Indians" OR Asian or "Asian Indian") AND ("United States")))
